# Data-Driven Predictive Modeling for Massive Intraoperative Blood Loss during Liver Transplantation: Integrating Machine Learning Techniques

**DOI:** 10.1101/2025.05.25.25328319

**Authors:** Taiichi Wakiya, Yukihiro Sanada, Noriki Okada, Yuta Hirata, Toshio Horiuchi, Takahiko Omameuda, Yasuharu Onishi, Yasunaru Sakuma, Hironori Yamaguchi, Yoshihiro Sasaki, Naohiro Sata

## Abstract

**Background:** Massive intraoperative bleeding (IBL) in liver transplantation (LT) poses serious risks and strains healthcare resources necessitating better predictive models for risk stratification. As traditional models often fail to capture the complex, non-linear patterns underlying bleeding risk, this study aimed to develop data-driven machine learning models for predicting massive IBL during LT using preoperative factors.

**Methods:** Two hundred ninety consecutive LT cases from a prospective database were analyzed. Logistic regression models were built using 73 preoperative demographic and laboratory variables to predict massive IBL (≥ 80 mL/kg). The dataset was randomly split (70% training, 30% testing). The model was trained and validated through three-fold cross-validation, with backward stepwise feature selection iterated 100 times across unique random splits. The final model, based on a high stability index, was evaluated using the area under the curve (AUC).

**Results:** Massive IBL was observed in 141 patients (48.6%). In standard logistic regression, significant differences were found in 42 of 73 factors between groups stratified by massive IBL, however, substantial multicollinearity limited interpretability. In the feature selection across 100 iterations, the data-driven model achieved an average AUC of 0.840 in the validation and 0.738 in the test datasets. The final model, based on 11 selected features with a high stability index, achieved an AUC of 0.844. An easy-to-use online risk calculator for massive IBL was developed and is available at: https://tai1wakiya.shinyapps.io/ldlt_bleeding_ml/.

**Conclusions:** Our findings highlight the potential of machine learning in capturing complex risk factor interactions for predicting massive IBL in LT.

## Introduction

Liver transplantation (LT) is a high-risk procedure frequently accompanied by substantial intraoperative bleeding (IBL), which can lead to adverse outcomes.(1–3) Additionally, perioperative bleeding and transfusions consume significant human and financial resources.(4, 5) Accordingly, effective strategies to anticipate and reduce bleeding are essential for optimizing outcomes and maintaining the sustainability of transplant programs.

Despite ongoing advances, predicting critical IBL during LT remains a major clinical challenge. Risk stratification tools using conventional statistical methods, such as logistic regression, have been developed to identify patients at increased risk.(6–9) However, these models often assume linearity and independence among variables, limiting their ability to capture the complex, multifactorial nature of LT-associated bleeding. As a result, their accuracy and clinical utility remain suboptimal.

Machine learning offers a promising alternative by allowing the integration of numerous interrelated factors without requiring pre-specified assumptions about variable relationships. ML models can capture nonlinear associations and detect complex patterns that traditional approaches or clinical intuition may overlook. This capability also extends to handling high-dimensional datasets and identifying subtle but clinically relevant trends, potentially offering novel insights into perioperative risk.(10, 11) This capability may enhance the predictive performance of bleeding models and facilitate more personalized perioperative management.

Although ML has shown value in various surgical contexts,(12–14) its application in predicting bleeding outcomes in LT is still limited. Therefore, we aimed to develop a data-driven, ML-based model using prospectively collected preoperative variables to predict massive IBL in LT. Our objective was to establish a clinically applicable framework that outperforms conventional models and supports individualized surgical planning.

## Materials and methods

### Patients

We conducted a retrospective monocentric observational study using a prospectively maintained database. This study was approved by the Ethics Committee of Jichi Medical University (Approval No. 20-008). This study was designed and conducted in accordance with the principles of the Declaration of Helsinki and Istanbul. The need for written informed consent for the present study was waived by the Institutional Review Board of Jichi Medical University in view of its retrospective design, in accordance with national and local guidelines, considering the fact that all clinical/laboratory measurements and procedures were part of routine care. This study included 290 consecutive patients who underwent living donor LT at our facility between 2008 and 2024.

### Surgical procedures and operative management

Donor hepatectomy was selected based on the recipient’s standard liver volume, weight, and graft volume determined using preoperative computed tomographic volumetry. For the recipient’s operation, inverted T-shaped or transverse incisions were made, and a total hepatectomy was performed. The graft hepatic vein was anastomosed to the stump of the recipient’s hepatic veins, which formed a single orifice, in an end-to-end manner. Hepatic artery reconstruction was routinely performed using microsurgical techniques. Choledocho-choledochostomy was the first choice for biliary reconstruction, except in cases in which the bile duct could not be used, such as with biliary atresia (BA) or primary sclerosing cholangitis.

### Definition of massive intraoperative bleeding

IBL was calculated on the basis of the in/out balance of the operative field. At our institution, fluid loss from the abdominal cavity, including that from bile and lymphatics, is considered IBL. IBL was divided by body weight and referred to as adjusted intraoperative bleeding (aIBL, mL/kg). Massive aIBL was defined as aIBL ≥ 80 mL/kg. In clinical practice, the estimated circulating blood volume is approximately 70–80 mL/kg.(15) Therefore, aIBL exceeding 80 mL/kg is effectively equivalent to the loss of the entire circulating blood volume, indicating a clinically significant hemorrhagic event. Accordingly, 80 mL/kg serves as a physiologically meaningful and practical cutoff.

### Data collection

For each patient, we collected demographic and laboratory data immediately before LT. The data were accessed for research purposes on April 22, 2025. Investigators had access to identifiable participant information during data collection; however, all data were de-identified prior to analysis. Seventy-three perioperative variables were extracted from the prospectively collected database. Patient demographic data included age, sex, body height, body weight, and etiology. Etiology was categorized as follows: acute liver failure (ALF), BA, graft failure, or other. Additionally, information regarding the history of previous abdominal surgery, ABO incompatibility, rituximab desensitization, and graft type was collected. Graft type was categorized as left lateral segment (LLS), left liver (LL), right liver (RL), right posterior segment (RPS), reduced LLS, or monosegment graft. To prevent statistical bias and enhance the generalizability of the model, incidence factors with a prevalence of less than 5%, such as spontaneous bacterial/fungal peritonitis and portal vein thrombosis, were excluded from the predictive variables. The 55 laboratory variables and their abbreviations are detailed in S1 Table. Prior to binary logistic regression analysis, the variables were standardized to have a mean of 0 and a standard deviation of 1, thereby ensuring that the logistic coefficients reflected the magnitude of influence on prediction. Given the study’s focus on identifying modifiable risk factors, particular emphasis was placed on preoperative blood test parameters, as these variables can be influenced by conditions such as infection and thrombosis and can be actively managed and optimized before surgery.

### Statistical analyses

Continuous variables were expressed as medians (ranges) and analyzed using nonparametric methods for non-normally distributed data (Mann–Whitney U test). Categorical variables were reported as numbers (percentages) and analyzed using the chi-square test or Fisher’s exact test, as appropriate. Variables with a significant relationship to massive IBL in univariate analysis were used in a binary logistic regression model. The correlation between the two parameters was analyzed using the Spearman rank-order method. Differences were considered significant at P < 0.05. Statistical analyses were performed using GraphPad Prism (v10.2.3; GraphPad Software, San Diego, CA; USA, https://www.graphpad.com).

### Data preparation and splitting

The overall workflow is illustrated using a block diagram in Fig 1. The dataset comprised 73 features along with corresponding binary labels. Data preprocessing involved a stratified random split, allocating 70% of the data for training and 30% for testing. To ensure a robust evaluation across different random splits, this process was repeated 100 times using unique random seeds (random_state = rr, where rr = 0, 1, …, 99).

**Figure 1:**
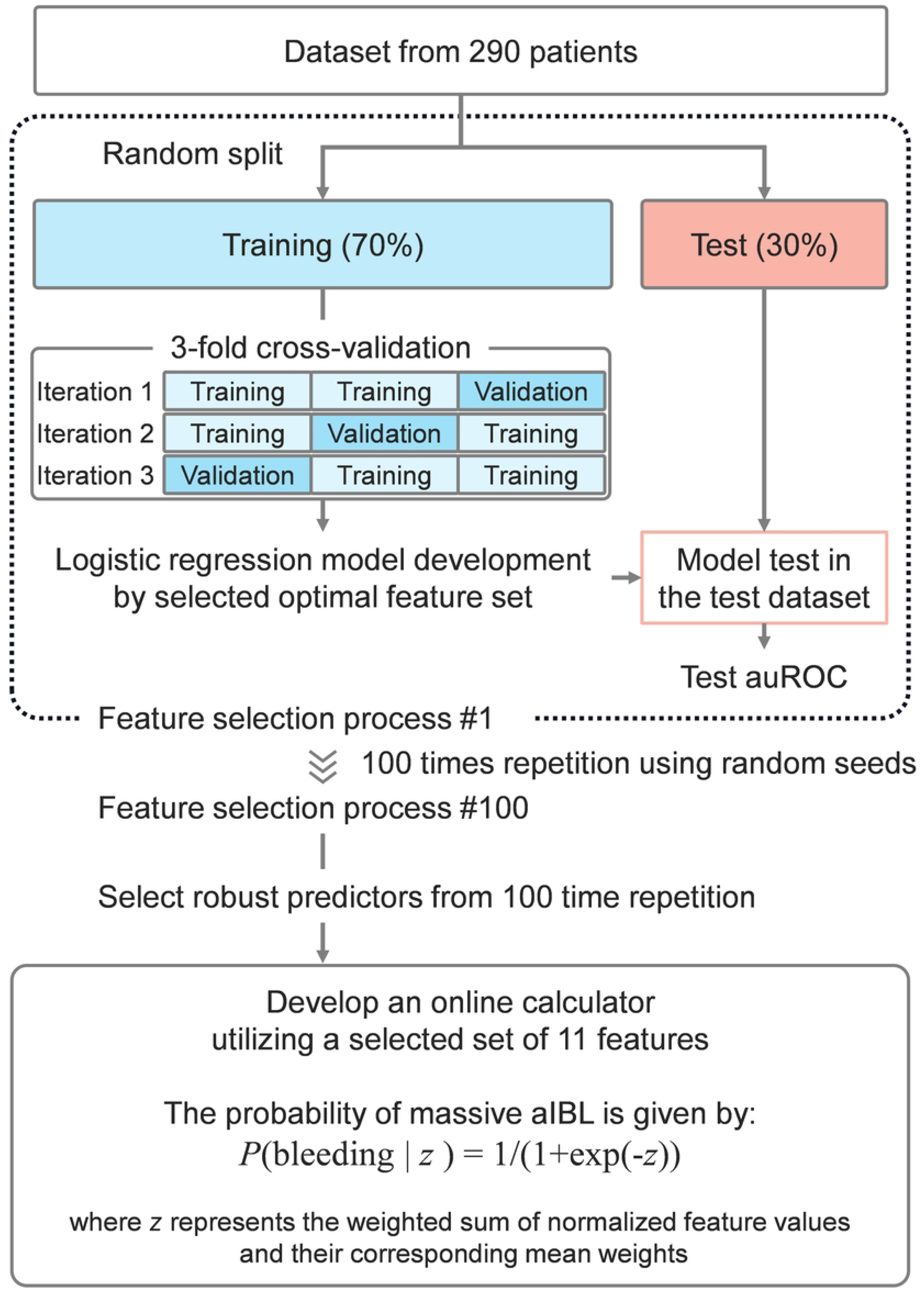
The study workflow and methodological process.

### Backward feature selection and model optimization

To identify the most informative predictors of massive intraoperative bleeding, we applied backward stepwise feature selection within a logistic regression framework. Model optimization and evaluation were conducted using stratified 3-fold cross-validation, repeated across 100 random data splits to ensure generalizability. At each iteration, variables were progressively removed based on their contribution to model performance, assessed using the area under the receiver operating characteristic curve (AUC). The feature set yielding the highest validation AUC was selected as the final model. Details of the feature elimination procedure, coefficient ranking, and model tuning are provided in the Supplementary Methods. All analyses were conducted using Python libraries including Scikit-learn and Pandas.(16)

### Development of an online calculator for estimating the risk of massive aIBL

We developed an online calculator based on the final logistic regression model to estimate the probability of massive aIBL. To ensure model robustness, feature stability was evaluated across multiple random data splits using occurrence rate and absolute mean weight. Features consistently selected with strong predictive weights were incorporated into the final model. Prior to application, selected features were standardized and weighted using their regression coefficients to calculate the predicted probability of massive aIBL. The calculator is available at: https://tai1wakiya.shinyapps.io/ldlt_bleeding_ml/. Details of the feature selection metrics, weighting procedures, and standardization process are provided in the Supplementary Methods.

## Results

### Clinical significance of massive aIBL

Among the 290 patients, those with aIBL greater than 80 mL/kg exhibited significantly higher rates of graft loss compared to those with lower aIBL (Fig 2). These findings support the clinical validity of using 80 mL/kg as a reasonable and actionable threshold for defining massive aIBL in LT.

**Figure 2:**
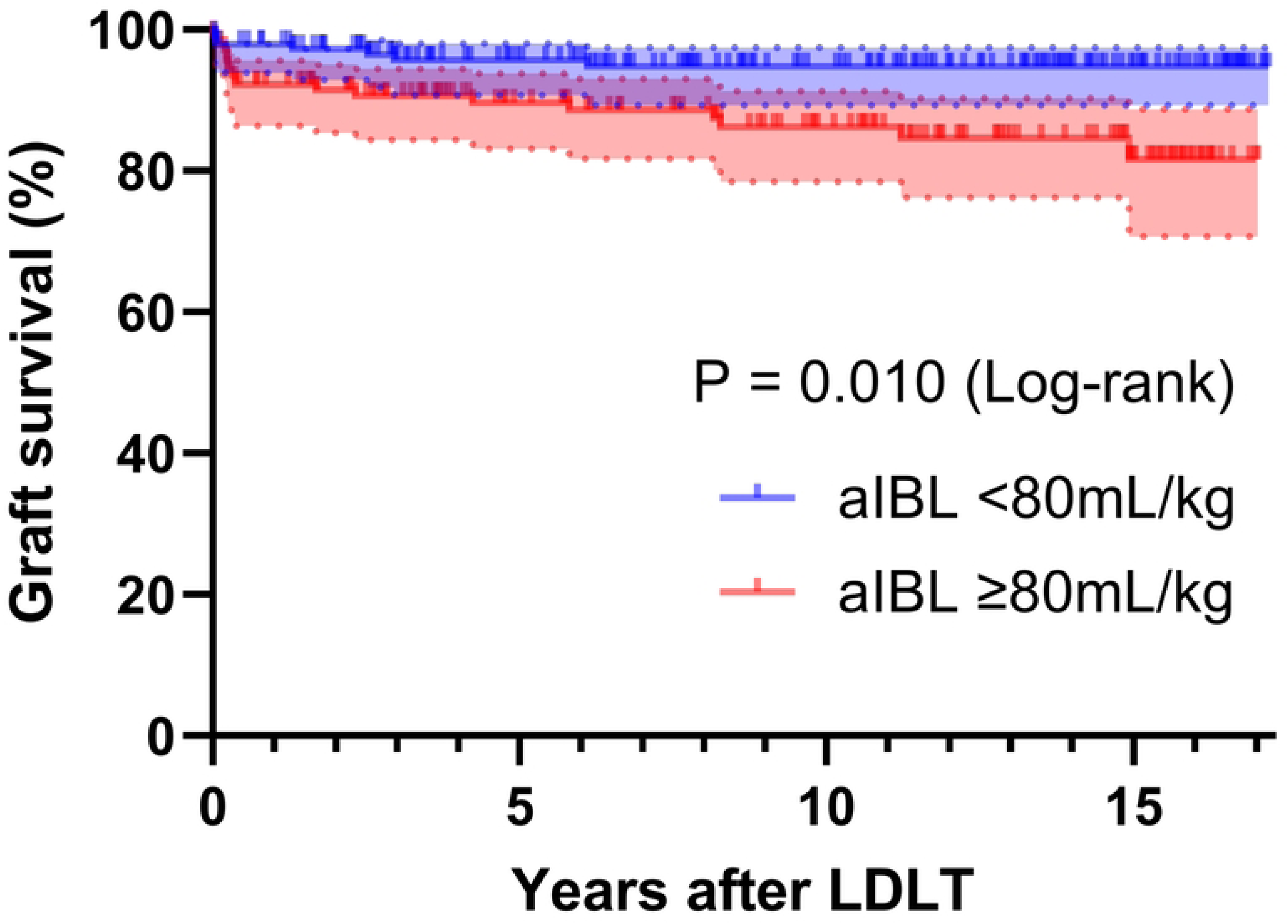
Graft survival curve stratified by adjusted intraoperative bleeding volume using an 80 mL/kg cutoff. IBL, intraoperative bleeding; LDLT, living donor liver transplantation.

### Comparison of patient characteristics in the massive aIBL and non-massive aIBL groups

The demographic data of the 290 enrolled patients are presented in Table 1, which also includes information on the IBL. Among all cases, massive aIBL was observed in 141 patients (48.6%). Except for indication and graft type, no significant differences were observed in the demographic data between the groups.

**Table 1.** Comparison of patient characteristics in the massive aIBL and non-massive.

|  | All<br>(n = 290) | aIBL<br><80mL/kg<br>(n = 149) | aIBL<br>≥80mL/kg<br>(n = 141) | P value |
| --- | --- | --- | --- | --- |
| Gender, female, n | 168 (57.9) | 88 (59.1) | 80 (56.7) | 0.778 |
| Age, year | 3 (0-69) | 3 (0-65) | 4 (0-69) | 0.606 |
| Body height, cm | 94.9 (46.0-178.0) | 95.6 (50.5-176.5) | 93.0 (46.0-178.0) | 0.480 |
| Body weight, kg | 14.2 (2.6-87.9) | 14.1 (2.6-72.5) | 14.4 (2.6-87.9) | 0.673 |
| Indication, n |  |  |  | 0.046 |
| ALF | 14 (4.8) | 3 (2.0) | 11 (7.8) |  |
| BA | 155 (53.5) | 83 (55.7) | 72 (51.1) |  |
| Graft failure | 13 (4.5) | 4 (2.7) | 9 (6.4) |  |
| Other | 108 (37.2) | 59 (39.6) | 49 (34.8) |  |
| History of previous abdominal surgery, n | 198 (68.3) | 98 (65.8) | 100 (70.9) | 0.415 |
| ABO incompatible, n | 58 (20.0) | 27 (18.1) | 31 (22.0) | 0.499 |
| Rituximab desensitization, n | 59 (20.3) | 34 (22.8) | 25 (17.7) | 0.352 |
| Graft type, n |  |  |  | <0.001 |
| LLS | 127 (43.8) | 78 (52.3) | 49 (34.8) |  |
| LL | 74 (25.5) | 43 (28.9) | 31 (22.0) |  |
| RL | 57 (19.7) | 20 (13.4) | 37 (26.2) |  |
| Monosegment | 16 (5.5) | 2 (1.3) | 14 (9.9) |  |
| Reduced LLS | 13 (4.5) | 5 (3.4) | 8 (5.7) |  |
| RPS | 3 (1.0) | 1 (0.7) | 2 (1.4) |  |
| IBL, mL | 1012 (30-64510) | 540 (30-5000) | 2603 (250-64510) | <0.001 |
| aIBL, mL/kg | 78.6 (1.6-1112.4) | 36.5 (1.6-79.5) | 159.5 (80.4-1112.4) | <0.001 |

aIBL, adjusted intraoperative bleeding; ALF, acute liver failure; BA, biliary atresia; IBL, intraoperative bleeding; LL, left liver; LLS, left lateral segment; RL, right liver; RPS, right posterior segment.

### Comparison of laboratory data in the massive aIBL and non-massive aIBL groups

Laboratory data are presented in Table 2. Significant differences (P < 0.05) were observed in 40 of the 55 laboratory test items between the two groups. To predict the occurrence of massive aIBL, we conducted a binary logistic regression analysis with the occurrence of massive aIBL as the dependent variable. Forty significant predictor variables associated with massive aIBL, identified using univariate analysis (P < 0.05), were included in the binary logistic regression analysis. The analysis identified total bile acids (P = 0.013, odds ratio [OR] = 1.010, 95% confidence interval [CI]: 1.000–1.010), albumin (P = 0.014, OR = 0.228, 95% CI: 0.071–0.736), and ammonia (P = 0.043, OR = 1.020, 95% CI: 1.000–1.050) as significant predictors of incidences of massive aIBL. However, as noted in S2 Table, there was significant multicollinearity among many factors in this analysis. The presence of multicollinearity can lead to instability in estimates, wider confidence intervals, and increased errors in coefficient estimates, complicating interpretation and potentially leading to incorrect conclusions. These results indicate that to construct a predictive model that exhibits multicollinearity with our dataset, it is crucial to select variables objectively, excluding subjectivity, and to utilize alternative analytical methods.

**Table 2.** Comparison of laboratory data in the massive aIBL and non-massive aIBL <u>groups</u>.

|  | All<br>(n = 290) | aIBL<br><80mL/kg<br>(n = 149) | aIBL<br>≥80mL/kg<br>(n = 141) | P value |
| --- | --- | --- | --- | --- |
| A2PI, % | 73.5 (16.4-150.0) | 86.1 (27.3-150.0) | 60.7 (16.4-116.1) | <0.001 |
| Alb, g/dL | 3.0 (1.7-4.6) | 3.2 (1.8-4.4) | 2.8 (1.7-4.6) | <0.001 |
| ALP, U/L | 803.0 (119.0-7793.0) | 857.0 (136.0-7452.0) | 688.0 (119.0-7793.0) | 0.320 |
| ALT, U/L | 49.5 (1.0-723.0) | 50.0 (1.0-467.0) | 48.0 (10.0-723.0) | 0.633 |
| AMY, U/L | 43.0 (3.0-387.0) | 45.0 (7.0-387.0) | 38.0 (3.0-320.0) | 0.027 |
| AnGap, mmol/L | 11.7 (2.0-26.4) | 12.0 (3.4-26.4) | 11.1 (2.0-19.6) | <0.001 |
| AST, U/L | 75.5 (14.0-1013.0) | 62.0 (16.0-565.0) | 88.0 (14.0-1013.0) | 0.148 |
| AT3, % | 67.1 (17.1-157.5) | 79.8 (21.8-157.5) | 47.3 (17.1-138.5) | <0.001 |
| BE, mmol/L | -1.2 (-9.5-12.0) | -1.4 (-9.5-8.9) | -0.7 (-8.0-12.0) | 0.063 |
| BS, mg/dL | 108.0 (51.0-349.0) | 111.0 (61.0-244.0) | 104.0 (51.0-349.0) | 0.024 |
| BUN, mg/dL | 8.0 (1.0-66.0) | 7.0 (1.0-66.0) | 9.0 (1.0-58.0) | <0.001 |
| Ca, mg/dL | 8.7 (7.4-12.4) | 8.8 (7.4-12.4) | 8.7 (7.5-12.4) | 0.006 |
| Ca2calc, mmol/L | 1.1 (0.7-1.4) | 1.1 (0.8-1.4) | 1.1 (0.7-1.4) | 0.417 |
| Che, U/L | 162.0 (25.0-443.0) | 176.0 (25.0-443.0) | 130.0 (33.0-436.0) | <0.001 |
| Cl, mmol/L | 107.0 (89.0-115.0) | 108.0 (90.0-115.0) | 106.0 (89.0-114.0) | <0.001 |
| CPK, U/L | 65.0 (10.0-981.0) | 67.0 (11.0-566.0) | 64.0 (10.0-981.0) | 0.624 |
| Crea, mg/dL | 0.24 (0.02-3.34) | 0.23 (0.02-1.79) | 0.24 (0.03-3.34) | 0.191 |
| CRP, mg/dL | 0.20 (0.01-7.59) | 0.13 (0.01-7.59) | 0.51 (0.01-6.81) | <0.001 |
| Dbil, mg/dL | 0.68 (0.01-39.06) | 0.28 (0.01-19.13) | 2.24 (0.01-39.06) | <0.001 |
| Ddimer, µg/mL | 1.1 (0.1-47.8) | 0.5 (0.1-16.6) | 2.4 (0.1-47.8) | <0.001 |
| Ferri, ng/mL | 42.6 (3.0-3526.1) | 28.3 (3.0-2370.0) | 61.3 (4.1-3526.1) | <0.001 |
| Fib, mg/dL | 173.5 (42.0-592.0) | 190.0 (42.0-592.0) | 166.0 (43.0-356.0) | 0.003 |
| GGT, U/L | 82.5 (8.0-1517.0) | 112.0 (8.0-1517.0) | 58.0 (8.0-1255.0) | 0.529 |
| Hb, g/dL | 9.1 (4.1-13.5) | 9.6 (6.4-13.5) | 8.6 (4.1-13.4) | <0.001 |
| HCO3 | 23.2 (16.8-36.5) | 22.8 (16.8-33.0) | 23.9 (17.6-36.5) | 0.004 |
| Hct, % | 27.1 (11.3-39.9) | 29.0 (19.8-39.9) | 25.7 (11.3-38.5) | <0.001 |
| K, mmol/L | 4.2 (2.1-8.0) | 4.1 (2.7-8.0) | 4.3 (2.1-6.2) | 0.340 |
| Lactate, mmol/L | 1.2 (0.4-10.4) | 1.2 (0.4-10.4) | 1.2 (0.5-8.5) | 0.549 |
| LDH, U/L | 219.0 (98.0-938.0) | 217.0 (98.0-938.0) | 230.0 (101.0-754.0) | 0.081 |
| MCH, pg | 29.7 (19.9-41.1) | 29.1 (19.9-39.1) | 30.7 (23.0-41.1) | <0.001 |
| MCHC, g/dL | 33.4 (30.0-36.1) | 33.4 (30.0-36.1) | 33.6 (30.3-36.0) | 0.036 |
| MCV, fL | 88.5 (65.0-118.6) | 86.0 (65.0-111.6) | 91.0 (72.6-118.6) | <0.001 |
| Mg, mg/dL | 2.0 (0.8-2.9) | 2.1 (1.3-2.5) | 2.0 (0.8-2.9) | 0.036 |
| Na, mmol/L | 139.0 (123.0-148.0) | 140.0 (123.0-148.0) | 138.0 (123.0-147.0) | <0.001 |
| NH3, μmol/L | 43.5 (4.0-161.0) | 39.0 (4.0-119.0) | 49.0 (15.0-161.0) | <0.001 |
| P, mg/dL | 4.1 (1.9-6.7) | 4.5 (2.0-6.4) | 3.8 (1.9-6.7) | <0.001 |
| PCO2, mmHg | 38.6 (28.3-102.0) | 37.6 (29.0-54.8) | 39.4 (28.3-102.0) | 0.003 |
| PH | 7.39 (7.13-7.54) | 7.40 (7.19-7.54) | 7.39 (7.13-7.53) | 0.887 |
| PIC, μg/mL | 0.5 (0.1-8.9) | 0.4 (0.1-2.9) | 0.6 (0.1-8.9) | <0.001 |
| Plasminogen, % | 62.1 (17.9-148.5) | 71.0 (23.2-148.5) | 54.2 (17.9-141.0) | <0.001 |
| Plt, $\times 10^4/\mu$L | 10.5 (1.4-66.5) | 12.6 (2.4-66.5) | 8.6 (1.4-34.9) | <0.001 |
| PO2, mmHg | 181.6 (69.4-463.0) | 191.0 (93.3-463.0) | 166.2 (69.4-386.3) | <0.001 |
| ProteinC, % | 44.5 (10.8-180.8) | 57.9 (12.0-180.8) | 34.0 (10.8-131.7) | <0.001 |
| PTINR | 1.35 (0.91-5.80) | 1.25 (0.91-2.72) | 1.47 (0.99-5.80) | <0.001 |
| PTpercent, % | 57.7 (8.8-115.1) | 68.4 (19.1-115.1) | 49.7 (8.8-95.6) | <0.001 |
| RBC, $\times 10^4/\mu$L | 310.5 (122.0-461.0) | 334.0 (210.0-461.0) | 278.0 (122.0-442.0) | <0.001 |
| TAT, ng/mL | 2.5 (0.5-218.1) | 2.0 (0.5-53.1) | 3.0 (0.6-218.1) | <0.001 |
| TBA, μmol/L | 67.2 (0.1-619.6) | 30.9 (0.1-493.6) | 119.5 (0.4-619.6) | <0.001 |
| Tbil, mg/dL | 2.74 (0.11-50.01) | 1.56 (0.11-25.63) | 6.18 (0.16-50.01) | <0.001 |
| Tchol, mg/dL | 142.0 (46.0-1999.0) | 149.0 (46.0-1999.0) | 134.0 (56.0-1115.0) | 0.010 |
| TCO2, mmol/L | 24.5 (17.8-38.3) | 23.9 (17.8-34.4) | 25.2 (18.6-38.3) | 0.002 |
| TG, mg/dL | 49.0 (10.0-370.0) | 49.0 (15.0-370.0) | 49.0 (10.0-342.0) | 0.499 |
| TP, g/dL | 5.7 (3.2-8.4) | 5.8 (4.3-7.3) | 5.6 (3.2-8.4) | 0.113 |
| UA, mg/dL | 4.0 (0.6-9.9) | 4.1 (0.7-9.5) | 3.9 (0.6-9.9) | 0.027 |
| WBC, $\times 10^3/\mu$L | 3.1 (0.5-15.7) | 3.4 (0.6-10.2) | 3.0 (0.5-15.7) | 0.251 |

aIBL, adjusted intraoperative bleeding; IBL, intraoperative bleeding.
Abbreviations for laboratory test items are detailed in S1 Table.

### Correlations between preoperative laboratory tests and intraoperative bleeding volume

Next, we examined the relationship between the preoperative laboratory test results and aIBL. Of the 55 parameters tested, 42 showed significant correlations with the bleeding volume. Positive correlations were found for 43.6% of the parameters, whereas 56.4% exhibited negative correlations. The top five positively correlated biomarkers were D-dimer (r = 0.588), prothrombin time international normalized ratio (r = 0.495), total bilirubin (r = 0.461), direct bilirubin (r = 0.438), and total bile acids (r = 0.363). Conversely, antithrombin III (r = -0.560), alpha 2-plasmin inhibitor (r = -0.555), protein C (r = -0.527), prothrombin time percent (r = -0.495), and plasminogen (r = -0.458) were highly negatively correlated (Fig 3). These findings suggest that these markers can potentially guide preoperative assessment and intervention.

**Figure 3:**
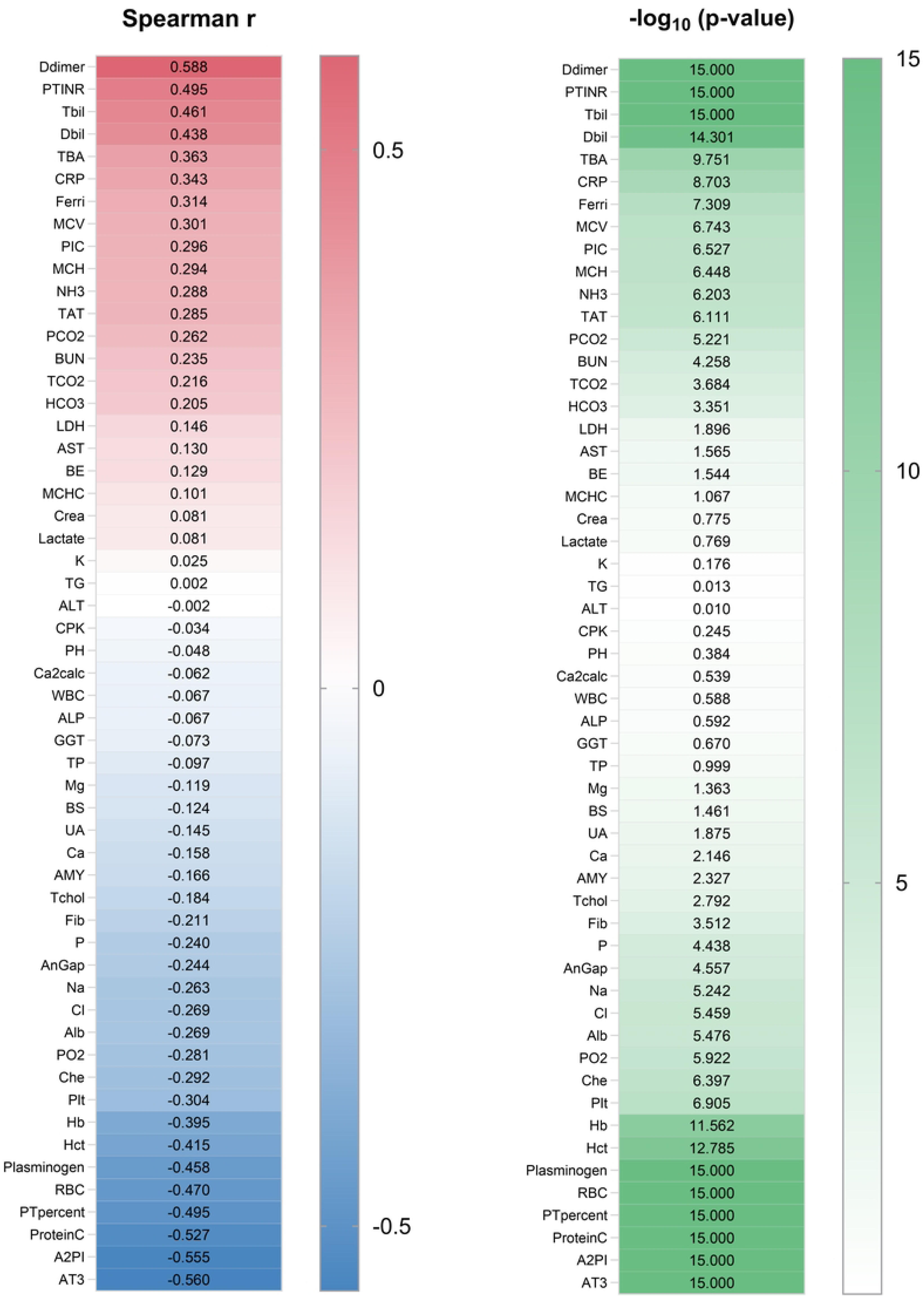
Correlations between preoperative laboratory tests and intraoperative bleeding volume. The left panel shows Spearman’s rank correlation coefficients, and the right panel shows p-values. All abbreviations and their units are listed in S1 Table.

### Feature selection and model performance in logistic regression for massive aIBL prediction

We further explored a more robust prediction model for massive aIBL. We used the backward stepwise method for feature selection to construct our binary logistic regression model. During this process, we calculated the absolute magnitudes of the coefficients and removed the least influential covariates. This analysis incorporated laboratory variables as well as demographic data, including age, categorical age (under 6 years, 6-18 years, or 18 years and older), sex, body height, body weight, etiologies (ALF, BA, graft failure, or other), history of previous abdominal surgery, ABO incompatibility, rituximab desensitization, and graft type (LLS, LL, RL, RPS, reduced LLS, or monosegment). We started with all 73 features and systematically removed the least significant ones from the set. One of the representative feature selection processes is presented in S3 Table and S1 Fig. The process for narrowing down the variables is detailed in S3 Table. Antithrombin III was identified as the final covariate in the dataset. S1 Fig illustrates the performance of the model at each stage. The highest validation AUC of 0.867 was observed at the stage with 12 covariates. The AUC obtained in the test dataset was 0.728. To ensure a robust evaluation, this process was repeated 100 times across different random splits (S2 Fig). The average number of selected features across these 100 trials was 15.1. The average AUC in the validation and test datasets was 0.840 (standard deviation [SD] = 0.022) and 0.738 (SD = 0.046), respectively. These findings support a systematic approach for preoperatively predicting massive IBL in LT, suggesting the robustness and reliability of feature selection and model performance.

### Identifying key predictors and constructing an online calculator for massive aIBL

To evaluate the consistency of selected features across 100 random data splits, index stability was assessed using occurrence rate and absolute mean weight (Fig 4). Based on the results from 100 iterations, features meeting the predefined criteria of an occurrence rate of at least 50% and an absolute mean weight of 0.5 or greater were selected. A total of 11 features were identified as robust predictors. Among them, six features exhibited positive mean weights, indicating a direct association with massive aIBL occurrence: D-dimer, ferritin, age, total bile acids, gamma-glutamyl transferase, and ammonia. Conversely, five features demonstrated negative mean weights, suggesting a potential protective effect: fibrinogen, platelet count, albumin, LLS graft, and antithrombin III. To enhance clinical applicability and ensure streamlined implementation, we developed a final prediction model for massive aIBL using these 11 selected features. In the test across all 290 cases, the model achieved an AUC of 0.844, with a sensitivity of 74.5%, specificity of 78.5%, positive predictive value of 76.6%, and negative predictive value of 76.5%. To further facilitate clinical adoption, we developed a user-friendly online calculator based on this risk model and made available online at: https://tai1wakiya.shinyapps.io/ldlt_bleeding_ml/.

**Figure 4:**
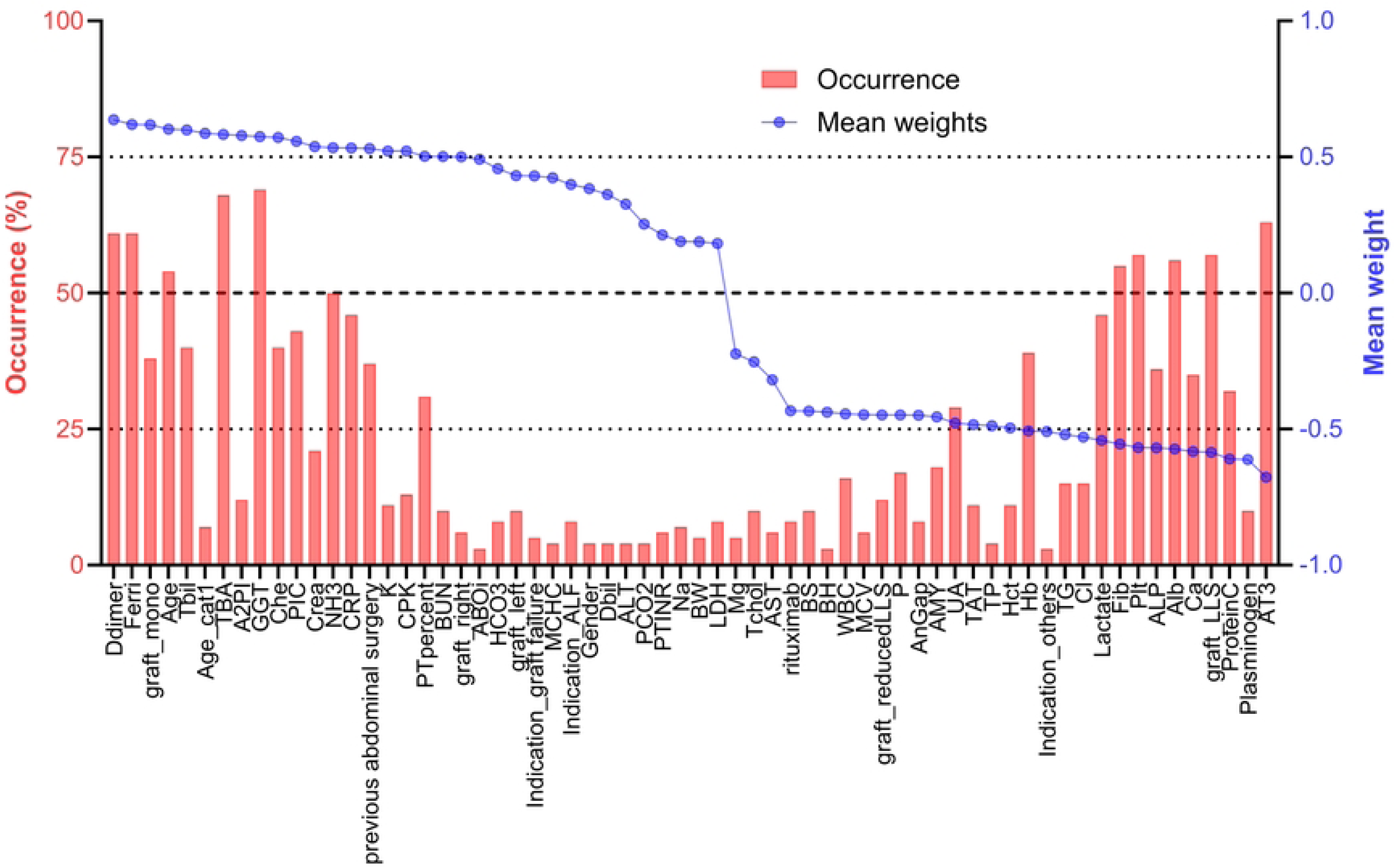
Index stability across 100 iterations. The index is arranged in order of occurrence, sorted by mean weight. All abbreviations and their units are listed in S1 Table.

## Discussion

In this study, we developed and validated ML models to predict massive IBL during LT. Using preoperative demographic and laboratory data, we employed logistic regression models with backward feature selection to achieve high predictive accuracy. Furthermore, we developed an online calculator that provides physicians with an accessible tool to estimate an individualized risk of massive IBL in the preoperative setting. To our knowledge, this is the first study to apply a ML model for IBL prediction in LT and to translate these findings into a clinically applicable prediction tool. The true innovation of this study lies not only in the excellent accuracy of the model but also in showcasing a groundbreaking artificial intelligence-based approach with significant future potential for management in LT.

Previous studies have attempted to predict bleeding and transfusion requirements in LT by using various clinical parameters and traditional standard statistical models.(6–9) While these studies have provided valuable insights, their models often exhibit suboptimal predictive performance and limited clinical applicability. Standard methods in clinical studies have several limitations, including the selection of variables, confounding factors, and multicollinearity, as demonstrated in this study. Even in prospective studies, the issue of multicollinearity remains a challenge to resolve. Furthermore, standard methods often fail to capture the complex, non-linear relationships between variables that significantly influence bleeding risk. In contrast, our study goes beyond traditional analyses by employing a data-driven approach, allowing us to systematically identify not only well-established risk factors but also previously unrecognized predictors.

Our study leveraged ML techniques, which are used to identify intricate patterns and interactions among multiple factors, demonstrating superior accuracy and robustness in predicting massive IBL. ML offers several key benefits, including increased accuracy through the simultaneous analysis of multiple variables and their interrelationships, and the ability to identify complex patterns and correlations that traditional methods might overlook. By incorporating a comprehensive set of preoperative variables and employing advanced feature selection methods, our ML models offer a more reliable and practical tool for preoperative risk stratification in LT. These models effectively manage complex, high-dimensional datasets, uncover hidden correlations, and explore non-linear medical relationships, enhancing predictive capabilities and potentially leading to new scientific insights.

Crucially, we went beyond model development by translating these findings into a tangible clinical tool. To ensure practical utility in real-world clinical settings, we developed an online calculator based on our predictive model, providing an accessible platform for physicians to rapidly assess massive IBL risk. This helps bridge the gap between advanced computational modeling and clinical decision-making, while integrating ML into clinical practice, which may contribute to improving patient outcomes and optimizing resource management.

Antithrombin III and ferritin emerged as significant predictors in our machine learning-based model, providing new insights into bleeding risk in LT. In cardiovascular surgery, low preoperative antithrombin III levels have been consistently associated with increased intraoperative blood loss.(17) This has led to the consideration of antithrombin III supplementation as a potential intervention. However, the outcomes of such supplementation remain inconclusive, with several studies and meta-analyses reporting inconsistent results.(18–21) Similarly, although the available studies are both dated and limited in number, evidence supporting a positive effect of antithrombin III supplementation in LT remains limited.(22, 23) Regarding ferritin, no direct evidence has previously established its role in predicting bleeding in liver surgery, including LT. Ferritin reflects both iron metabolism and systemic inflammation, and its elevation may indicate a preoperative inflammatory state that predisposes patients to coagulopathy.(24) Inflammatory responses are known to disrupt hemostasis through mechanisms such as endothelial dysfunction, hyperfibrinolysis, and altered coagulation cascades.(25) Supporting this, our analysis identified a strong positive correlation between IBL and C-reactive protein, another established inflammatory marker; however, ferritin demonstrated greater stability as a predictive index than C-reactive protein. Together, these findings illustrate how machine learning can reveal biologically and clinically relevant predictors that are often overlooked by conventional approaches. Antithrombin III and ferritin may serve as valuable biomarkers for risk stratification and perioperative management in LT.

The findings of this current study should be interpreted in light of several limitations. First, it was a single-institution cohort study with a relatively small patient population. Additional training data could potentially enhance the prediction accuracy. Furthermore, the lack of external validation using an independent dataset is also a limitation. One reason for the absence of external validation is the argument made by some biostatistics experts in predictive research stating that independent verification can be misleading and should be omitted as a model evaluation step.(26, 27) These experts report that simulations confirming at least 100 events and 100 non-events are required for a reliable assessment of predictive performance. They suggest using all available data for model development, with some form of cross-validation or bootstrap validation to assess the statistical optimism in average predictive performance.(27, 28) Based on these biostatistical perspectives, we chose to build our ML model using all data with cross-validation, performing 100 iterations with randomly split datasets to ensure robustness and generalizability. However, we acknowledge that the results of predictive research with small sample sizes are exploratory in nature.(27) Nevertheless, external validation in various clinical settings, covering the heterogeneity among cases, is essential for clinical application. Given the high prediction accuracy of our method, further development using large databases, such as national or regional datasets, is expected and necessary.

## Conclusions

Our study demonstrates the potential of ML to predict massive IBL during LT. These data-driven predictive models could revolutionize preoperative planning and intraoperative management, ultimately improving patient care and resource efficiency. Future research should focus on external validation using large datasets and the integration of these models into clinical practice.

## Data Availability

Data cannot be shared publicly because of institutional policy and patient confidentiality. Data are available from the Jichi Medical University Institutional Ethics Committee (contact via https://www.jichi.ac.jp/kenkyushien/clinical/clinical_human/) for researchers who meet the criteria for access to confidential data.

## Author contribution

T.W. contributed to the study conception and design. T.W., Yu.S., N.O., Y.H., T.H., T.O., Y.O., Ya.S., H.Y., and N.S. collected the clinical data. T.W. and Yo.S. analyzed the data. T.W. wrote the first draft of the manuscript paper. Yu.S., N.O., Y.H., T.H., T.O., Y.O., Ya.S., H.Y., Yo.S., and N.S. contributed to the review, and/or critical revision of the manuscript. All authors have approved the final article.

## Ethics statement

This study was approved by the Ethics Committee of Jichi Medical University (Ethics Committee Approval Case Number 20-008). Informed consent was obtained in the form of opt-out on our website (https://www.jichi.ac.jp/transplant/contents/disclosure.html), with the approval of the Ethics Committee of Jichi Medical University.

## Data availability statement

The data used in this study are not publicly available due to institutional policy and patient confidentiality. As participant consent for public sharing was not obtained, the data cannot be shared openly. However, anonymized data may be available from the corresponding author (TW) upon reasonable request.

## Figure Legends

**Supplemental Figure 1:** Representative performance of the binary logistic regression model at each stage of the backward stepwise feature selection process. This figure illustrates a single iteration out of 100 repetitions. AUC, area under the receiver operating characteristic curve.

**Supplemental Figure 2:** Model performance evaluation based on a feature selection process repeated 100 times. This figure presents the most accurate outcome observed among the selection processes conducted in each random split. AUC, area under the receiver operating characteristic curve.

## References

1. Rana A, Petrowsky H, Hong JC, Agopian VG, Kaldas FM, Farmer D, et al. Blood transfusion requirement during liver transplantation is an important risk factor for mortality. J Am Coll Surg. 2013;216(5):902–7.

2. Yuasa T, Niwa N, Kimura S, Tsuji H, Yurugi K, Egawa H, et al. Intraoperative blood loss during living donor liver transplantation: an analysis of 635 recipients at a single center. Transfusion. 2005;45(6):879–84.

3. Ramos E, Dalmau A, Sabate A, Lama C, Llado L, Figueras J, et al. Intraoperative red blood cell transfusion in liver transplantation: influence on patient outcome, prediction of requirements, and measures to reduce them. Liver Transpl. 2003;9(12):1320–7.

4. Stokes EA, Wordsworth S, Staves J, Mundy N, Skelly J, Radford K, et al. Accurate costs of blood transfusion: a microcosting of administering blood products in the United Kingdom National Health Service. Transfusion. 2018;58(4):846–53.

5. Ruiz J, Dugan A, Davenport DL, Gedaly R. Blood transfusion is a critical determinant of resource utilization and total hospital cost in liver transplantation. Clin Transplant. 2018;32(2).

6. Priem F, Karakiewicz PI, McCormack M, Thibeault L, Massicotte L. Validation of 5 models predicting transfusion, bleeding, and mortality in liver transplantation: an observational cohort study. HPB. 2022;24(8):1305–15.

7. Giabicani M, Joly P, Sigaut S, Timsit C, Devauchelle P, Dondero F, et al. Predictive Role of Hepatic Venous Pressure Gradient in Bleeding Events Among Cirrhotic Patients Undergoing Orthotopic Liver Transplantation. JHEP Reports. 2024:101051.

8. Thibeault F, Plourde G, Fellouah M, Ziegler D, Carrier FM. Preoperative fibrinogen level and blood transfusions in liver transplantation: A systematic review. Transplantation Reviews. 2023;37(4):100797.

9. Pustavoitau A, Rizkalla NA, Perlstein B, Ariyo P, Latif A, Villamayor AJ, et al. Validation of predictive models identifying patients at risk for massive transfusion during liver transplantation and their potential impact on blood bank resource utilization. Transfusion. 2020;60(11):2565–80.

10. Ray S, editor A Quick Review of Machine Learning Algorithms. 2019 International Conference on Machine Learning, Big Data, Cloud and Parallel Computing (COMITCon); 2019 14-16 Feb. 2019.

11. Xu Y, Liu X, Cao X, Huang C, Liu E, Qian S, et al. Artificial intelligence: A powerful paradigm for scientific research. The Innovation. 2021;2(4):100179.

12. Banerjee A, Dashtban A, Chen S, Pasea L, Thygesen JH, Fatemifar G, et al. Identifying subtypes of heart failure from three electronic health record sources with machine learning: an external, prognostic, and genetic validation study. The Lancet Digital health. 2023;5(6):e370–e9.

13. Taleb I, Kyriakopoulos CP, Fong R, Ijaz N, Demertzis Z, Sideris K, et al. Machine Learning Multicenter Risk Model to Predict Right Ventricular Failure After Mechanical Circulatory Support: The STOP-RVF Score. JAMA cardiology. 2024;9(3):272–82.

14. Wakiya T, Ishido K, Kimura N, Nagase H, Kubota S, Fujita H, et al. Prediction of massive bleeding in pancreatic surgery based on preoperative patient characteristics using a decision tree. PLOS ONE. 2021;16(11):e0259682.

15. Wilson F. Blood volume maintenance and restoration. In: Burton VW, Davies AH, Kilpatrick A, McIllmurray MB, Pring JE, Wilson F, editors. Essential Accident and Emergency Care. Dordrecht: Springer Netherlands; 1981. p. 130-40.

16. Pedregosa F. Scikit-learn : Machine learning in python. J Machine Learn Res. 2011;12:2825–30.

17. Laveson JE, Winford J, Iden J, Marengo-Rowe AJ, Race GJ. Investigation Of The Relationship Between Preoperative Chromogenic Antithrombin III Assays And Hemorrhage Following Heart Surgery. Thromb Haemost. 1981;46(05):0937.

18. Paparella D, Rotunno C, De Palo M, Finamore S, Guida P, Rubino G, et al. Antithrombin Administration in Patients With Low Antithrombin Values After Cardiac Surgery: A Randomized Controlled Trial. The Annals of Thoracic Surgery. 2014;97(4):1207–13.

19. Jooste EH, Scholl R, Wu Y-H, Jaquiss RDB, Lodge AJ, Ames WA, et al. Double-Blind, Randomized, Placebo-Controlled Trial Comparing the Effects of Antithrombin Versus Placebo on the Coagulation System in Infants with Low Antithrombin Undergoing Congenital Cardiac Surgery. Journal of Cardiothoracic and Vascular Anesthesia. 2019;33(2):396–402.

20. Slaughter TF, Mark JB, El-Moalem H, Hayward KA, Hilton AK, Hodgins LP, et al. Hemostatic effects of antithrombin III supplementation during cardiac surgery: results of a prospective randomized investigation. Blood Coagulation & Fibrinolysis. 2001;12(1):25–31.

21. Li T, Bo F, Meng X, Wang D, Ma J, Dai Z. The effect of perioperative antithrombin supplementation on blood conservation and postoperative complications after cardiopulmonary bypass surgery: A systematic review, meta-analysis and trial sequential analysis. Heliyon. 2023;9(11):e22266.

22. Baudo F, DeGasperi A, deCataldo F, Caimi TM, Cattaneo D, Redaelli R, et al. Antithrombin III supplementation during orthotopic liver transplantation in cirrhotic patients: a randomized trial. Thrombosis research. 1992;68(4-5):409–16.

23. Palareti G, Legnani C, Maccaferri M, Gozzetti G, Mazziotti A, Martinelli G, et al. Coagulation and fibrinolysis in orthotopic liver transplantation: role of the recipient’s disease and use of antithrombin III concentrates. S. Orsola Working Group on Liver Transplantation. Haemostasis. 1991;21(2):68–76.

24. Kell DB, Pretorius E. Serum ferritin is an important inflammatory disease marker, as it is mainly a leakage product from damaged cells. Metallomics. 2014;6(4):748–73.

25. Iba T, Helms J, Neal MD, Levy JH. Mechanisms and management of the coagulopathy of trauma and sepsis: trauma-induced coagulopathy, sepsis-induced coagulopathy, and disseminated intravascular coagulation. Journal of thrombosis and haemostasis : JTH. 2023;21(12):3360–70.

26. Steyerberg EW, Harrell FE, Jr. Prediction models need appropriate internal, internal-external, and external validation. Journal of clinical epidemiology. 2016;69:245–7.

27. Steyerberg EW. Validation in prediction research: the waste by data splitting. Journal of clinical epidemiology. 2018;103:131–3.

28. Steyerberg EW, Harrell FE, Jr., Borsboom GJ, Eijkemans MJ, Vergouwe Y, Habbema JD. Internal validation of predictive models: efficiency of some procedures for logistic regression analysis. Journal of clinical epidemiology. 2001;54(8):774–81.

